# Non-invasive SARS-CoV-2 genome surveillance and its utility in resource-poor settings during the Delta wave of the COVID-19 pandemic

**DOI:** 10.1101/2023.02.16.23286031

**Authors:** Krishna Khairnar, Siddharth Singh Tomar

**Affiliations:** Environmental Virology Cell (EVC), Council of Scientific and Industrial Research-National Environmental Engineering Research Institute (CSIR-NEERI), Nagpur, India; Academy of Scientific and Innovative Research (AcSIR), Ghaziabad, UP, India

**Author notes:** Corresponding author: Correspondence to Krishna Khairnar.

## Abstract

Voluntary participation of the public in disease surveillance can be encouraged by deploying user-friendly sample collection processes that can minimise the discomfort to the participants. This study evaluated the suitability of saline gargle-based sample collection for genomic surveillance of SARS-CoV-2, which included 589 SARS-CoV-2 positive samples collected from Nagpur city in central India from March to December 2021. The SARS-CoV-2 positive samples were subjected to whole genome sequencing (WGS) using the oxford nanopore technologies next-generation sequencing platform. Out of 589 samples, 500 samples qualified for the WGS, and the results revealed eight different clades of SARS-CoV-2 encompassing 37 different Pango-lineage types. The mutation mapping analysis of the variants identified in this study showed six mutations of interest and one mutation of concern E484K in the spike glycoprotein region. Our findings indicate that non-invasive gargle-based genomic surveillance is scalable and does not need significant changes to the existing workflow post-sample collection.

## 1. INTRODUCTION

Severe acute respiratory syndrome coronavirus 2 is an RNA virus that causes the coronavirus disease of 2019 or COVID-19. The SARS-CoV-2 virus first appeared in Wuhan, China, in December 2019, unleashing a pandemic that is still going strong today.[1,2] As of February 14, 2023, the number of people infected by the SARS-CoV-2 virus is more than 755 million, and the total number of deaths is above 6.8 million [3]. These figures clearly show how harmful and highly transmissible this virus is. The virus’s genome significantly influences the pathogenicity of the virus. The SARS-CoV-2 genome is approximately 30 kb long and encodes 26 proteins [4]. Any changes in the nucleotide sequences in the genome can result in mutations that can change the amino acid encoded and, subsequently, the protein produced. This will result in the formation of new virus variants [5]. The significant variants of concern identified till now are Alpha, Beta, Gamma, Delta, and Omicron [6] The rate of mutation affects the transmissibility and infectivity of the virus considerably. These adaptive mutations of the virus can lead to the formation of new variants which may be resistant to therapy and vaccines, thus making the control of the pandemic difficult. Therefore, timely diagnosis and genomic surveillance are essential to monitoring potentially harmful viral variants worldwide. Wastewater surveillance is being hailed as an indirect and non-invasive method for SARS-CoV-2 genome surveillance as it covers a large area and does not require in-person sampling. Several reports showed that wastewater surveillance gives an early warning of emerging cases in a given area [7,8,9]. Wastewater surveillance provides a preliminary picture of the disease spread by estimating the presence or absence of the disease in a particular area but can not resolve the source from where the disease originated.

Wastewater surveillance may be useful, especially in many regions that lack sufficient resources for deploying large-scale molecular testing platforms for public health monitoring. An emerging viral transmission in the population may be indicated if a wastewater sample shows the SARS-CoV-2 virus in a previously reported low-prevalence area. The authorities may use this information to issue warnings or take administrative measures. Hence, knowing the viral presence in wastewater can give indicative information about the location/site of an initial circulation, which can be resolved further by directing the resources to the location/site to trace infected cases. However, there are concerns about the sensitivity and reliability of wastewater surveillance methods, as wastewater is a highly diluted and complex system. Seasonal variations affect the composition and concentration of wastewater, making it a very inconsistent system to work on. Detecting viruses in the wastewater system depends on several aspects, like the wastewater network, structure, capacity, virus-shedding profile, wastewater properties, sampling strategy, methodologies, and detection limit [10]. As the virus is released into wastewater, it travels through a long and complex wastewater network until it reaches the location from where the sample is collected. In the underdeveloped and developing world, the integrity of wastewater systems is questionable. The wastewater streams from industrial, domestic, grey-water and stormwater drains are often mixed due to compromised channels that are not well segregated [11]; such mixing may adversely affect the quality of viral detection. The wastewater travel time is affected by various factors, including the branching and structure of the wastewater network and the flow rate during the sampling period.

Considering the limitations of wastewater monitoring, the situation warrants an alternative approach to ensure reliable genome surveillance in the post-pandemic scenario that can be achieved by the voluntary participation of the public. The nasopharyngeal-oropharyngeal swab (NPS-OPS) based molecular testing, and genome surveillance is the existing gold standard but cause considerable discomfort to persons due to invasive sample collection [12]; also, improper NPS-OPS sample collection due to an unskilled technician can lead to misleading results [13], which poses a limitation in promoting voluntary participation of the public. A non-invasive and patient-friendly sampling method could find more acceptance amongst the public for voluntary participation during monitoring. Non-invasive sampling methods such as saliva and gargling could be useful in post-pandemic surveillance. It is also important to note that saliva and gargle-based sampling methods can efficiently recover the virus from the oropharynx and buccal area. Still, the nasopharynx site remains unsampled in these methods, which may be a limitation. Nevertheless, such non-invasive sampling methods hold the potential for large-scale monitoring.

Therefore, to increase sample collection throughput and reduce patient discomfort, the Council of Scientific and Industrial Research-National Environmental Engineering Research Institute (CSIR-NEERI) developed a non-invasive, patient-friendly saline gargle sample collection method for diagnosing SARS-CoV-2. This method can also be deployed for sample collection of other respiratory viruses. The saline gargle sample collection method was approved to be used in India by the Indian Council of Medical Research (ICMR) [14]. The Drugs Controller General of India (DCGI) approved the Industrial scaleup and kit manufacturing of the saline gargle method [15].

The saline gargle method has several advantages over conventional swab-based sample collection methods. This method is ideal for self-collection as it is non-invasive and does not require trained healthcare professionals for sample collection. The saline-gargle kit includes a tube containing 5 ml of phosphate-buffered saline solution. The user must gargle with this solution for 15 seconds, followed by 15 seconds of rinsing in the mouth and then releasing the wash in the tube with the help of a disposable collection funnel and sealing it with a screw-capped cap. The saline gargle effectively rinses and collects the virus from the buccal cavity and the oropharynx. The sample collected through this method is relatively free from polymerase chain reaction (PCR) inhibitors which is the case with viral transport medium (VTM) swab-based sampling techniques. NPS-OPS are uncomfortable for patients; during the sample collection, the irritation may lead to sneezing and coughing reflexes which may risk direct exposure to healthcare personnel and surrounding [12].

The saline gargle method also circumvents the need for the time-consuming and costly RNA extraction procedure; the viral RNA from the sample can be isolated by using a one-step RNA release buffer. The RNA release buffer is added to the collected saline gargle sample and incubated at room temperature for 30 minutes, followed by heat inactivation at 98 degrees Celsius for 6 minutes to isolate an RNA suitable for further use as a template for PCR and Whole Genome Sequencing (WGS).

Voluntary participation of the public in disease surveillance can be encouraged by deploying user-friendly sample collection processes that minimise the discomfort to the participants. The simplicity and scalability of Gargle-based sample collection also make it an ideal candidate suitable for post-pandemic disease surveillance. Although the quantitative reverse transcriptase real-time PCR (qRT-PCR) is the gold standard for SARS-CoV-2 detection, the qRT-PCR can not resolve between the variants of SARS-CoV-2. Therefore, the WGS-based genome surveillance of SARS-CoV-2 is required for variant-level identification. Setting up sequencing-based genomic surveillance in low-income nations can aid in the collection of accurate surveillance data as well as help in long-term outbreak management. However, it concerns what is practically possible in that specific region or environment. For WGS, the extracted RNA is converted into cDNA and analysed using next-generation sequencing (NGS) techniques. Therefore, the yield and quality of extracted RNA are crucial for getting quality sequencing results. Our study evaluated the suitability of saline gargle-based sample collection for genomic surveillance of SARS-CoV-2. This study included 589 SARS-CoV-2 positive samples collected using the Gargle-based sample collection method from Nagpur city in central India from March to December 2021. The study focused on the utility of a patient-friendly sample collection method for remote, resource-poor, and undeveloped parts of the world to perform genome surveillance of respiratory viruses like SARS-CoV-2. Also, the study gives projections for the utility of deploying patient-friendly, fast, and economical sample collection strategies such as saline gargling for post-pandemic surveillance.

## 2. MATERIALS AND METHODS

### 2.1 Data collection

The metadata of 589 SARS-CoV-2-positive patients from March to December 2021 was taken from the Integrated Health Information Platform (IHIP) [16]. The metadata of SARS-CoV-2 positive cases includes patient details such as age, gender, qRT-PCR cycle threshold (Ct) value, and SARS-CoV-2 variant information. The information on symptoms and vaccination coverage of the patients was sought from the Indian council of medical research (ICMR) COVID-19 data portal; 42.5% of patients reported being symptomatic, and 57.4% were found to be asymptomatic. 13.4% of patients were vaccinated with Covaxin, 82.5% were vaccinated with the Covishield vaccine, 0.7% with the Sputnik V vaccine and vaccine information was unknown for 3.1% of the patients.

### 2.2 Materials

The SARS-CoV-2 viral RNA was extracted using an RNA release buffer containing Tris-EDTA and Proteinase K. Applied Biosystems QuantStudio 3, and StepOnePlus qRT-PCR machines were utilised for qRT-PCR, cDNA synthesis, PCR tiling, and rapid barcoding. Qubit 4 Fluorometer by Thermo Fisher Scientific was used to quantify nucleic acids for quality control (QC) of DNA libraries for Oxford Nanopore Technology (ONT)-based WGS. SARS-CoV-2 WGS was done on an Mk1C 6.3.9 and Mk1B ONT MinION sequencing platform.

### 2.3 Sample collection and RNA extraction

The saline gargle samples were collected as a part of a novel genome surveillance initiative launched by the Nagpur Municipal Corporation (NMC) in collaboration with CSIR-NEERI, Nagpur, for the city of Nagpur from March to December 2021. Nagpur is the third largest city in the Indian state of Maharashtra and the fourteenth largest city in India by population [17]. 589 SARS-CoV-2 positive saline gargle samples were selected for the genome surveillance study. Overall gender distribution in the sample set was 54.8% males and 45% females. In this study, the percentage distribution of the cases within the age groups was 0.4% (0 to <2 years), 1.4% (2 to <5 years), 4% (5 to <15 years), 59% (15 to <50 years), 23% (50 to <65 years), and 12% (≥ 65 years). The age distribution was according to the WHO Global Epidemiological Surveillance Standards for Influenza [18]. The sample collection was carried out using a saline gargle collection kit, and the viral RNA was isolated using an RNA release buffer. The standard operating procedure developed by CSIR-NEERI for Saline gargle-based sample collection and one-step RNA isolation for SARS-CoV-2 detection by RTPCR and WGS is available [19]. A documentary on CSIR-NEERI’s Saline Gargle-based SARS-CoV-2 RT-PCR detection is also available [20]. The isolated RNA samples were used immediately or stored at −80°C till further use.

### 2.4 RT-PCR for detecting SARS-CoV-2

qRT-PCR was performed for quantitative detection of SARS-CoV-2 using MBPCR255 Hi-PCR COVID-19 Triplex Probe PCR Kit (HIMEDIA) or Meril SARS-CoV-2 kit in each sample. The primer-probe sets in the kits specifically detect viral RNA from SARS-CoV-2. The samples with a Ct value of ≤ 38 were considered RT-PCR positive.

### 2.5 SARS-CoV-2 whole genome sequencing

The isolated RNA samples of 589 SARS-CoV-2 positive samples were selected for SARS-CoV-2 WGS. The cDNA synthesis was done using the TAKARA Prime Script RT reagent kit (RR037A) [21]. The sequencing libraries were constructed by multiplex PCR tiling according to the protocol of ONT [22]. The prepared cDNA libraries were sequenced using MinION Mk1C or MinION Mk1B.

### 2.6 Bioinformatic analysis

The live base-calling was performed using the Guppyv22.10.7 base-calling algorithm integrated into the MinION Mk1C sequencer [23]. The processed FASTQ reads from the MinION sequencer were analysed using the bioinformatics platform COMMANDER developed by Genotypic Technology Pvt. Ltd [24]. COMMANDER is a graphical user interface software developed to ease bioinformatic analysis post-sequencing. The FASTA sequence and the variant call performed by the COMMANDER pipeline were further confirmed by the web-based Pangolin COVID-19 Lineage Assigner [25]. The metadata, including the FASTA sequence, was submitted to the Global initiative on sharing Avian Influenza Data (GISAID) and the Indian biological data centre (IBDC). The mutation mapping of SARS-CoV-2 variants was done using the Outbreak.info web server [26]. Outbreak.info server calculates the prevalence of mutations as a ratio of the number of sequences carrying a given set of mutations on a given day at a specific place (or all locations) (x) divided by the total number of sequences on that day in that location (n).

## 3. RESULTS & DISCUSSION

This study has attempted to evaluate the utility of the gargle-based sample collection method for SARS-CoV-2 genome surveillance. Ct values of qRT-PCR are inversely proportional to the viral load in the sample. The Ct value range of samples analysed in this study was from 10 to 38 (**Fig.1**). The gargle-based sample collection was able to detect the virus by RT-PCR in cases with lesser viral load; an earlier study with 250 paired samples of saline-gargle and VTM-swab from patients revealed that gargle sample was able to detect the virus in samples with lesser viral load (data unpublished). The gargle-based sample collection also enabled the successful WGS of the positive samples with lesser viral loads. Generally, for a reasonably good WGS read, the Ct value of the positive sample must be ≤25. However, the gargle-based genome surveillance has enabled the WGS of positive samples even with a Ct value above 35, which has a lesser viral load.

**Figure 1:**
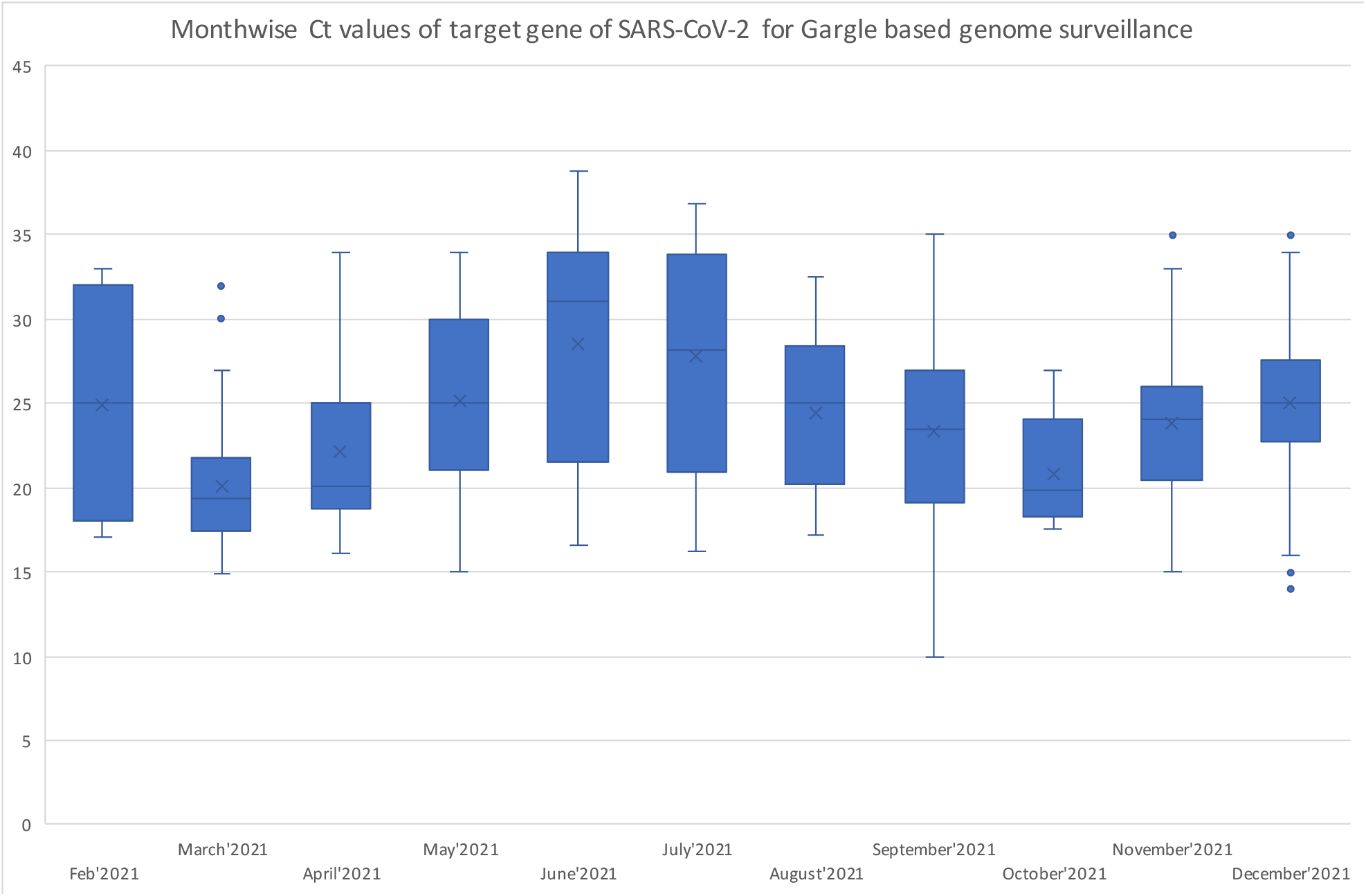
Monthly Ct values of the target gene of SARS-CoV-2 for Gargle-based genome surveillance from March to December 2021

Of 589 samples, only 500 qualified for the SARS-CoV-2 WGS variant calling by enforcing the criteria of ≥70% genome coverage and sequencing coverage of ≥100X. WGS result of the SARS-CoV-2 positive samples revealed a diverse variant profile comprising 37 different Pango-lineage types categorised into eight different clades of SARS-CoV-2 (**Fig 2**). The SARS-CoV-2 variants which had a percentage share of ≥1% in the study were AY.112 (38.8%), B.1.617.2 (15.6%), B.1 (6.6%), AY.127(6.4%), B.1.617.1 (5.2%), AY.43 (5%), AY.122 (2.8%), AY.100 (2.4%), AY.120 (2%), AY.112.2 (1.8%), AY.102 (1.8%), AY.75 (1.8%), AY.39 (1.4%), AY.16 (1.2%), and AY.65 (1.2%). The eight clades, as per the next clade, had a respective share of 21J (68.4%), 21A (17.2%), 20A (6.8%), 21B (5.2%), 21B (1.8%), 21K (0.20%), 20B (0.20%) and 20I (0.20%) (**Fig 2**). Overall variant percentage share among the samples and variants showing <1% percentage share is classified as other variants represented in supplementary **FigureS1**. 21J (Delta) is a variant of concern (VOC) Delta. This clade has spread in Europe, the Americas, Africa, and Oceania. 21J(Delta) carries all mutations of 21A along with mutation at position G215C. Additionally, other amino-acid mutations at ORF1a:A1306S, ORF1b:V2930L, ORF1a:T3255I, ORF1a:T3646A, ORF1b:A1918V, and ORF7b:T40I were reported in 21J clade. Clade 21A was first detected in India and had spike mutations L452R and P681 impacting antibody binding. 21A has some additional mutations in spike protein at positions T19R, R158G, T478K and D950N.

**Figure 2:**
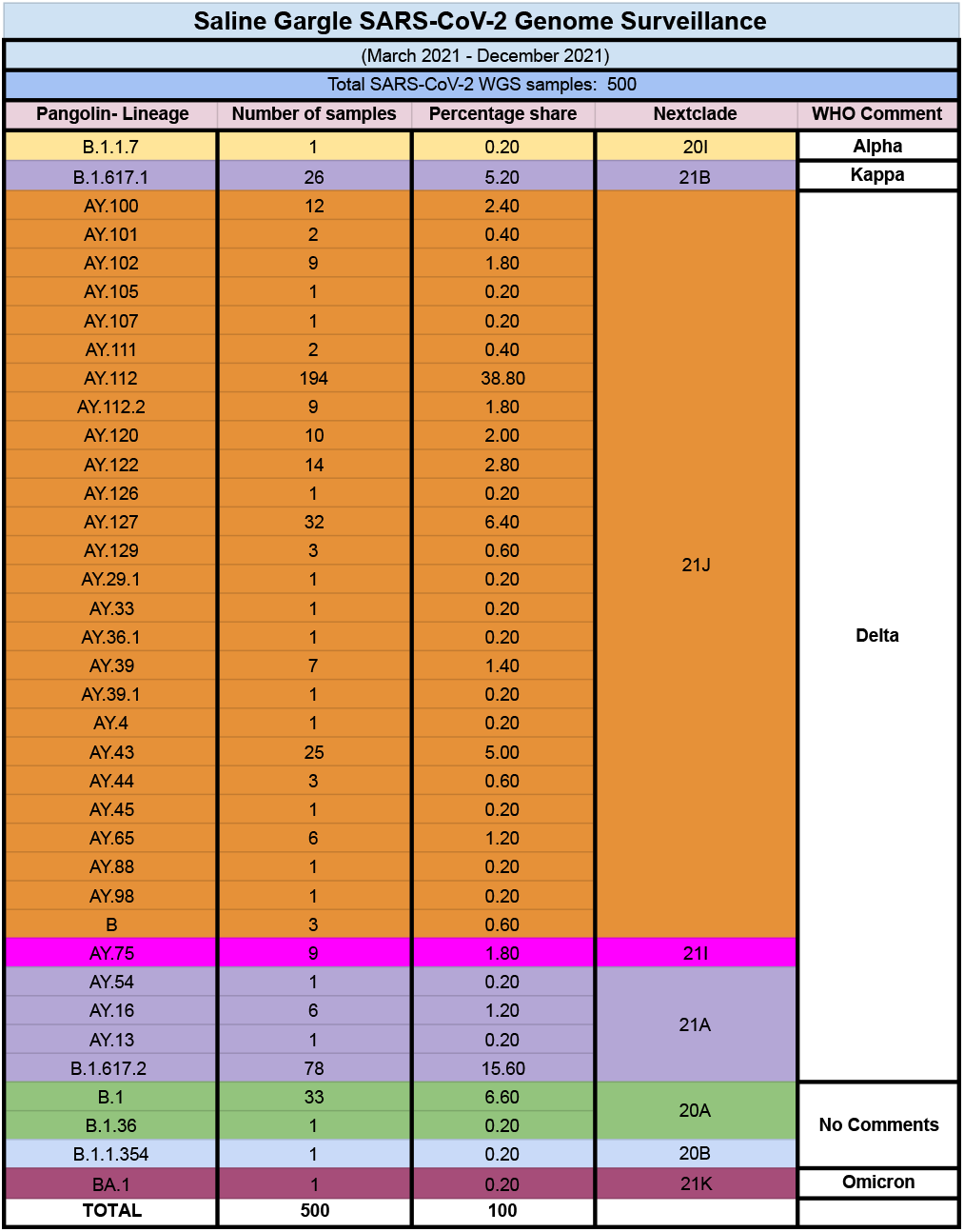
Summary of variants and their corresponding clades found in the SARS-CoV-2 WGS of saline gargle samples from March to December 2021

In this study, variant AY.112 appeared as an overall dominating variant. It was also noted that AY.112 consistently appeared with a percentage share of at least >15% throughout the period from March to December 2021. AY.112 showed a maximum prevalence (63.2%) in September. An interesting trend was observed in this study concerning AY.112, which started emerging in March and progressed to appear competitively till June with B.1.617.2 and completely replaced B.1.617.2 in July. However, B.1.617.2 reemerged in August and continued to prevail till December. Another variant, AY.127, which first emerged in August, continued to increase from October to December, showing >27% percentage share in December. A detailed month-wise variant distribution of variants and relative percentage share is summarised in **Figures 3a and 3b**.

**Figure 3a:**
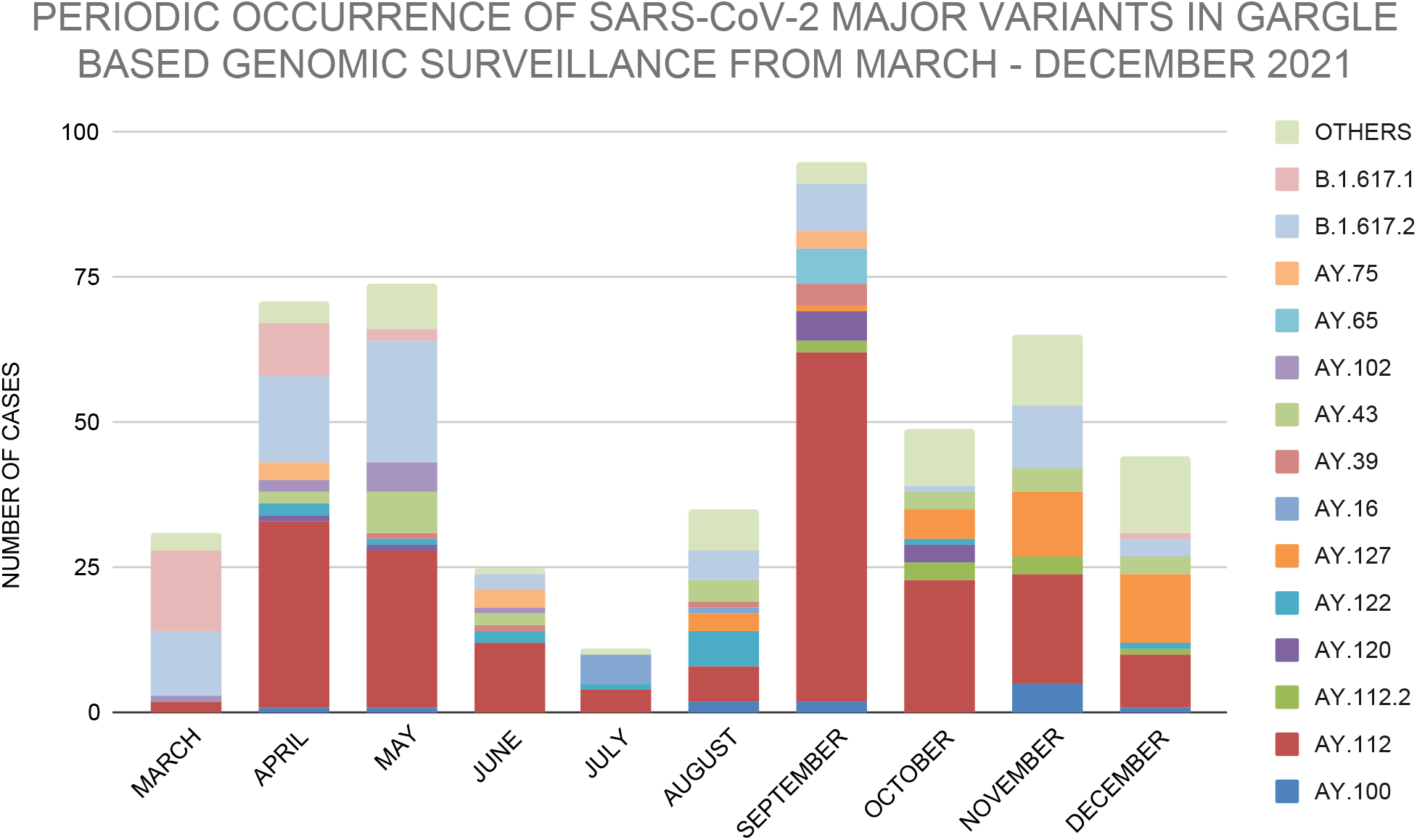
Monthly occurrence of SARS-CoV-2 major variants in gargle-based genomic surveillance from March to December 2021

**Figure 3b:**
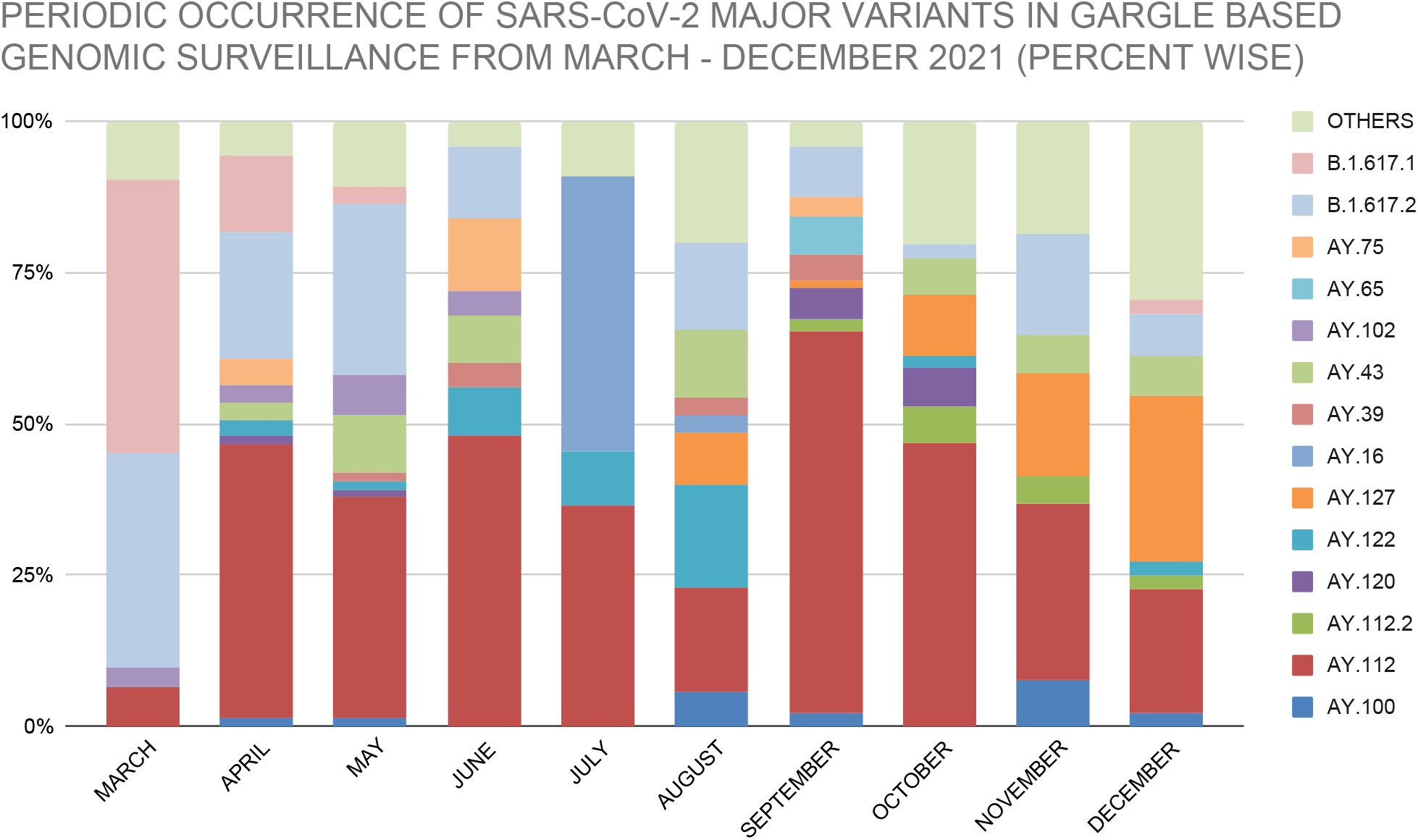
Percentage-wise monthly occurrence of SARS-CoV-2 major variants in gargle-based genomic surveillance from March to December 2021

Thirty-seven different variants were detected in the study, and these variants were then subjected to Mutation mapping analysis using the outbreak.info web server. The results showed Six mutations of interest (MOI), including K417N, L452R, S477N, N501Y, P681H, P681R, and one mutation of concern (MOC) E484K in the spike glycoprotein region, as shown in **Figure 4**. The MOI and MOC percentage prevalence of all variants identified in the study among the GISAID sequences has been summarised in **Figure 5**. Briefly, the K417N mutation was found to be 0.5-1 % MOI prevalence among the GISAID sequences of all samples except the BA.1 variant of Omicron lineage in which K417N mutation showed 99% of MOI prevalence; The Mutation L452R appeared with 90-100% MOI prevalence in the majority of the variants belonging to kappa and delta lineages, except in B.1 (5%), B.1.36 (0.5%), B.1.1.354 (7.7%), BA.1 (0.5%) and B.1.1.7 (Alpha) (0.5%) MOI prevalence. The S477N mutation was found to be 0.5-0.8 % MOI prevalence across the studied variants except for the BA.1, where the mutation S477N appeared with 88% of MOI prevalence. The N501Y mutation showed 0.5-4% MOI prevalence in most of the studied variants except B.1.1.7 and BA.1, which showed 98% and 84% MOI prevalence, respectively. P681H showed 0.5-1% MOI prevalence among the majority of variants and showed 99% MOI prevalence in B.1.1.7 and 98% MOI prevalence in BA.1. However, mutation P681R appeared in the majority of variants from delta lineages with >95% MOI prevalence interestingly P681R showed no prevalence among AY.122, AY.126, AY.29.1, AY.33, B.1.36, and B.1.1.354, and a meagre prevalence of <0.5% among B.1.1.7, B.1, and BA.1. The MOC E484K remained undetected in B.1.617.1, AY.101, AY.102, AY.111, AY.112.2, AY.29.1, AY.39, AY.65, AY.88, AY.98, AY.16 and AY.13 and showed <0.5% prevalence among the rest of the studied variants.

**Figure 4:**
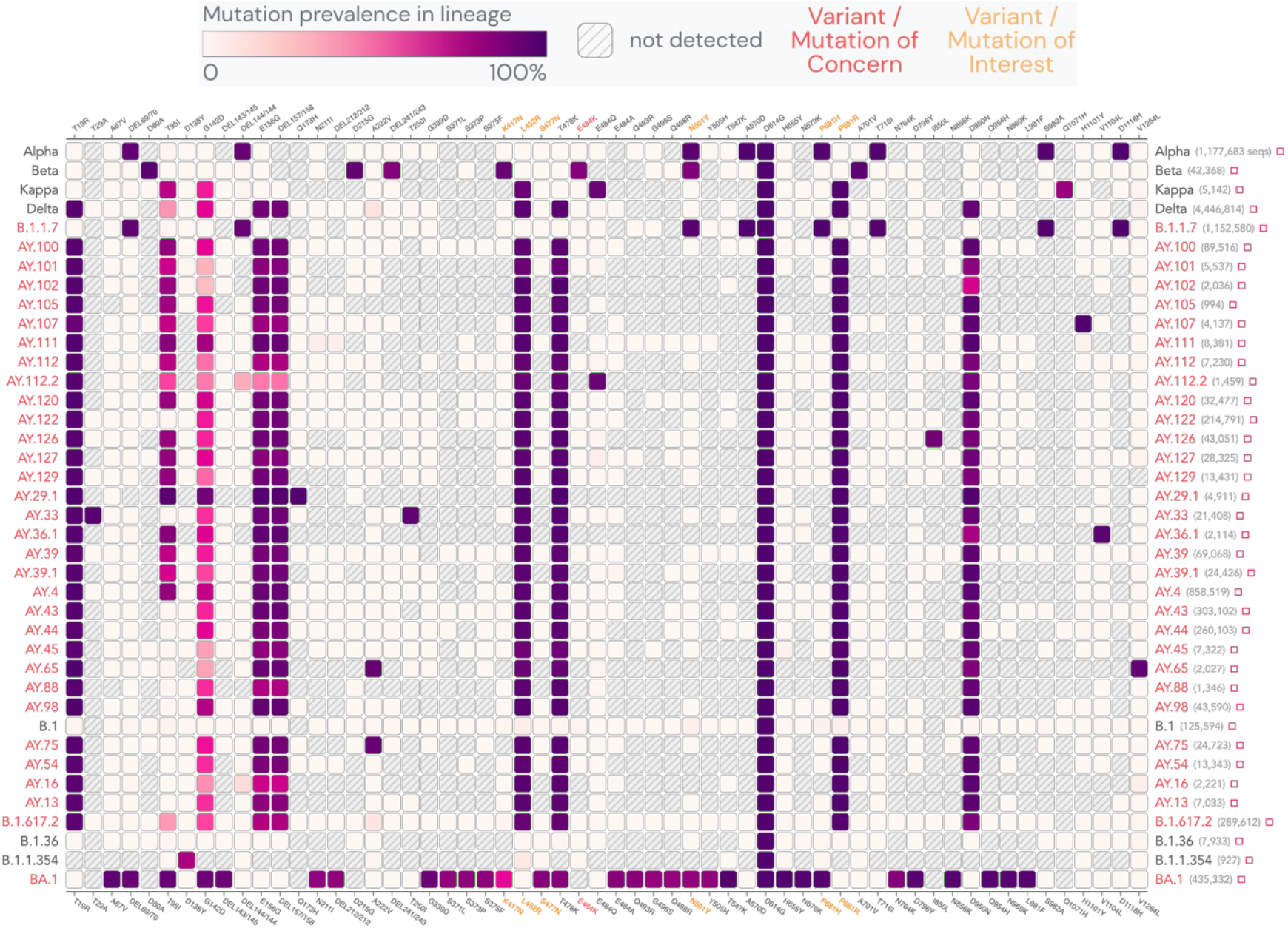
Spike glycoprotein gene mutation mapping of epidemiologically significant SARS-CoV-2 variants identified in the study from March to December 2021.

**Figure 5:**
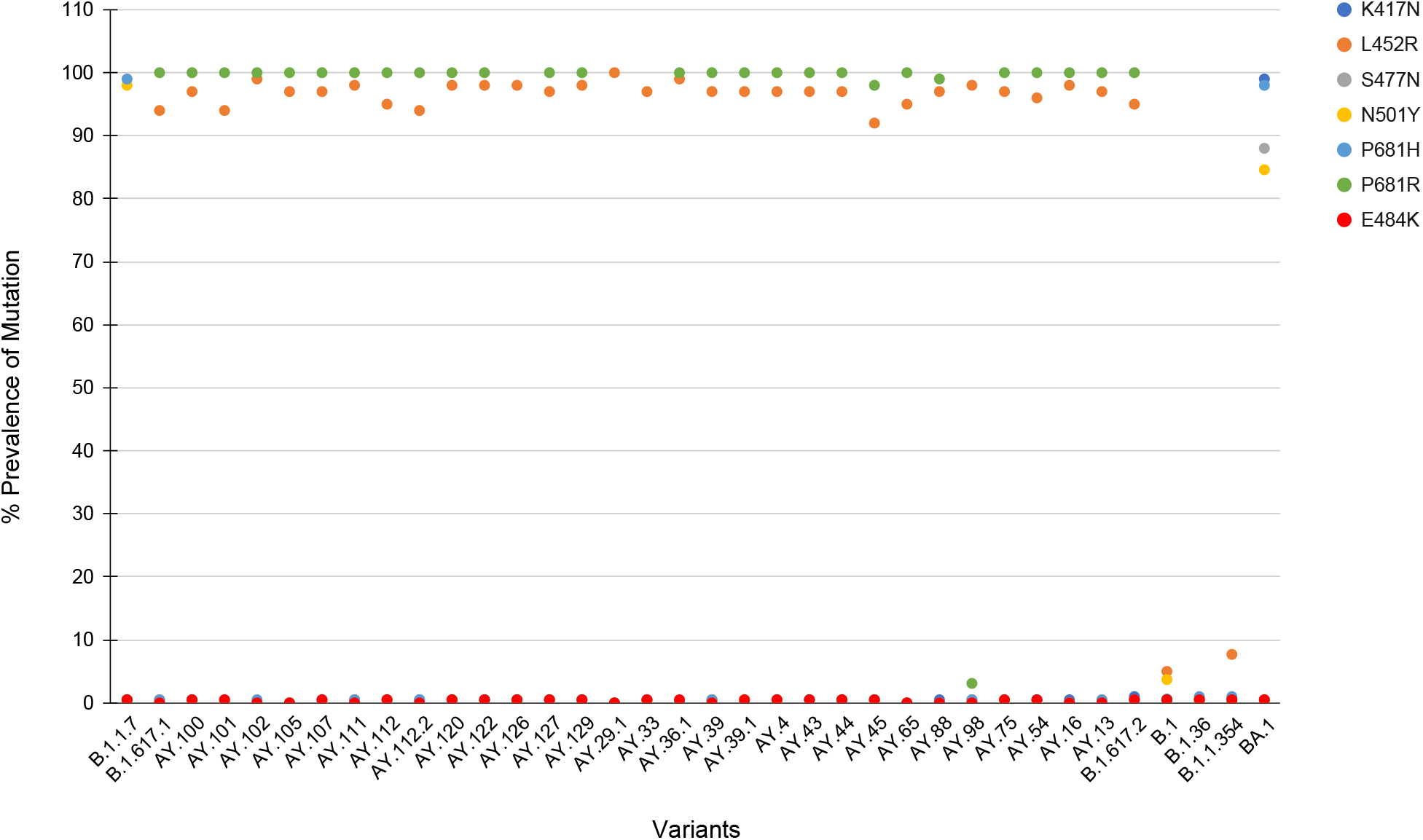
Spike glycoprotein gene mutation of interest and mutation of concern percentage prevalence among the GISAID sequences for all variants identified in the study.

The SARS-CoV-2 variants having L452R mutation are reported to have a decreased susceptibility to convalescent and vaccinated sera and mAbs. [27] According to genome-wide phylogenetic analysis, acquiring the L452R mutation could be responsible for the emergence of both Kappa and Delta variants. Global analysis of SARS-CoV-2 sequences showed that L452R was present in various independently emerging SARS-CoV-2 B.1.617 variants [27]. The SARS-CoV-2 Delta variant becoming a globally dominant variant could also be attributed to the P681R mutation leading to Delta’s replacement of the Alpha variant during the pandemic. Delta P681R mutation enhances the cleavage of the full-length spike to S1 and S2 subunits while interacting with the ACE2 receptor, which could improve cell-surface-mediated virus entry [28]. Studies demonstrated that the Delta variant exhibited improved infectivity and reduced susceptibility to vaccine-induced neutralising antibodies compared to the wild-type Wuhan-Hu. When tested invitro, the spike proteins of Wuhan-Hu (P681) and BA.2 (H681) pseudoviruses showed enhanced cell fusion and syncytia formation, and Delta spike (P681R) demonstrated improved fusogenic activity and syncytia formation capabilities. Live-viruses plaque formation assays confirmed these findings and demonstrated that relative to the wild-type, Delta formed more plaques [29].

Genomic surveillance for respiratory viruses should be a regular protocol for early detection of any new outbreak, However, this is not the case; it can be observed that genomic surveillance performed is concentrated in some parts of the world, while in other parts, no data is being generated. Only 45 countries of the world, accounting for only 38%, perform high-quality genomic surveillance, while 17 countries perform moderate levels of sequencing and 31 countries perform limited genomic surveillance. About 76 countries worldwide do not provide any genomic sequencing data for surveillance [30,31]. In many underdeveloped or low-income countries, the collection and transport of samples, lack of facilities, the overall cost of sequencing, etc., can lead to such outcomes. To counter the emergence of novel viral variants and proper outbreak surveillance, appropriate techniques for sample collection, sample processing, and sequencing techniques suitable for undeveloped regions of the world are required. Therefore, collecting, processing, and sequencing samples from even the most remote geographic locations and rural communities is imperative. This study has projections for post-pandemic surveillance by deploying patient-friendly, rapid, and economical sample collection strategies. The type of sample used and the collection technique can also significantly impact the sequencing results. The WHO lists NPS-OPS samples, bronchoalveolar lavage, sputum, saliva, gargles, mouthwashes, etc., as samples with high viral RNA content and could potentially be used for genomic surveillance [32]. Among these, NPS are the most commonly used specimens for diagnosis and genomic surveillance; hence considered the gold standard collection technique. After collection, these NPS-OPS swabs are placed in a VTM and sent for RNA extraction, followed by screening RT-PCR. Collecting NPS-OPS samples requires technical expertise, which may or may not be available elsewhere.

It is also not practical to deploy experts to remote locations because it would be technically challenging to frequently collect sufficient samples for diagnosis and sequencing. Improper sample collection is also a significant problem using NPS [13]. Patient non-cooperation due to physical discomfort often makes it difficult for even experienced personnel to collect these samples accurately. It is important to note that the primary function of VTM had been transportation to ensure the viability and stability of the virus for the culture of viral samples. However, we must consider the fact that very few VTM samples sent for testing and sequencing are getting cultured. Thus, whether VTM is necessary for testing and genome sequencing arises. The ability of gargle-based genome surveillance to yield quality WGS even in lower viral load samples could be due to a lack of PCR inhibitors in the gargle medium. The phosphate-buffered saline (PBS) holds lesser interfering PCR inhibitors.

On the other hand, the routine VTM used in the NPS-OPS-based sample collection is reported to contain specific PCR inhibitors [30]. The antibiotics present in the VTM to inhibit bacterial growth may degrade the bacterial cells and release intracellular enzymes such as proteases and nucleases in the medium; these enzymes could adversely impact PCR kinetics by compromising the nucleic acid template if the cold chain is broken while transportation [34]. Kirkland et al. (2020) showed that commercially produced VTM solutions negatively influence the capacity to identify SARS-CoV-2 and Influenza virus RNA. A study on the RNA extracted from samples collected using commercial VTMs revealed that VTM components interfere with the PCR kinetics during PCR while amplifying viral RNA in these samples. This study used Phosphate buffered saline (PBS) added with 0.5% gelatin as a reference VTM and compared commercially available VTMs with it; the findings showed that the RNA was stable in the PBS-Gelatin media for 48 hours at room temperature, while no RNA was detectable in commercial VTMs after 48 hours at room temperature [35]. The transport of NPS-OPS in VTM must be done under the cold chain. There is a looming risk of RNA degradation due to a break in the cold chain while it is transported from resource-poor locations like remote or rural areas to distantly located testing and sequencing facilities. The lack of infrastructure and technical expertise in the rural and remote parts of the world can often lead to the loss of samples that could otherwise be used for generating important surveillance data. Such shortfalls need to be addressed by exploring alternatives.

The genomic surveillance process starts with sample collection, and ensuring a good quality sample is crucial for successful genomic surveillance. Conventional sample collection methods for respiratory viruses such as NPS-OPS, Bronchoalveolar lavage, and anterior nares swabs are invasive techniques and, thus, discourage the voluntary participation of the public in genomic surveillance. The saline gargle sample collection is non-invasive and patient-friendly, which will be more acceptable to the public. Gargle-based sample collection is a self-collection method and does not require any healthcare worker for sample collection; therefore, its scalability in resource-poor settings is better than the conventional methods. For post-pandemic monitoring, policymakers are focusing on employing waste-water genomic surveillance; however, monitoring the outbreaks in remote and rural areas and areas not connected to any wastewater network is not feasible for sustained genomic surveillance. The concept of wastewater-based SARS-CoV-2 monitoring is highly biased towards an urban set-up, which completely ignores the rural set-up. We believe that the non-invasive approaches like the saline gargle method have an advantage over the wastewater surveillance method for post-pandemic monitoring, especially in developing countries like India, where a very large proportion of humanity (65%) still lives in rural areas [36] which are not connected to any wastewater system. A large population in the urban area of India is also not connected to a wastewater system. According to the 2011 Census of India, 38% of urban households, that is, 30 million homes, rely on stand-alone septic tanks which are not connected to any wastewater network [37]. In a situation like this, entirely relying on the wastewater-based surveillance technique may not be a good proposition. It is important to reassess the utility of wastewater-based genome surveillance and consider alternative approaches. Therefore, gargle-based genomic surveillance is scalable and does not require significant deviations from the workflow post-sample collection, making it suitable for post-pandemic surveillance applications. The advantage of the saline gargle technique is the ease of sample collection and user-friendliness; patients can collect the samples independently without requiring qualified technicians. The mouth rinses or gargle-based sample collection method offers comparable results to the commonly used swab-VTM-based sample collection method for SARS-CoV-2 detection [38,39,40]. SARS-CoV-2 is a respiratory virus, so the gargle collection technique yields similar results to a throat swab [41]. The gargle collection technique covers a larger surface in the buccal area and the throat, offering a better sample consortium. This non-invasive sample collection technique can be particularly advantageous for underdeveloped or remote areas as a reliable sample collection technique for public surveillance in post-pandemic scenarios, which can find more user acceptance than the invasive swab-VTM sample collection method.

## Data Availability

All data produced in the present study are available upon reasonable request to the authors

---

**Figure S1:**
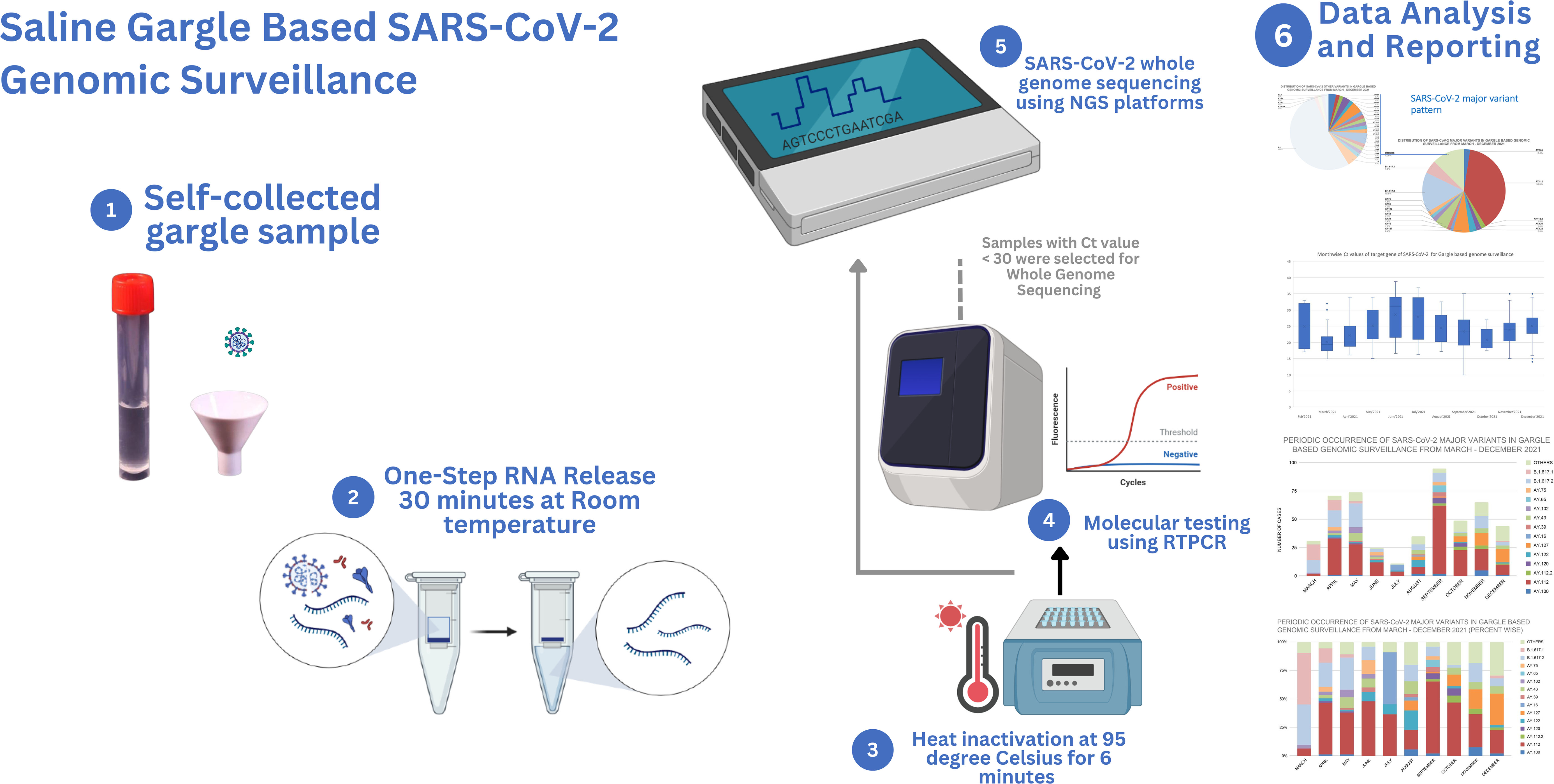
Overall share of SARS-CoV-2 variants identified by saline gargle-based WGS for SARS-CoV-2

## ACKNOWLEDGMENTS

This work was supported and funded by CSIR-NEERI, Nagpur. Some graphics in the graphical abstract have been taken from Biorender. It is certified that the manuscript has been checked for plagiarism by the institute knowledge resource centre through iThenticate (anti-plagiarism software) KRC No. CSIR-NEERI/KRC/2023/FEB/EVC/2.

## REFERENCES

1. Zhu, N., Zhang, D., Wang, W., Li, X., Yang, B., Song, J., Zhao, X., Huang, B., Shi, W., Lu, R., Niu, P., Zhan, F., Ma, X., Wang, D., Xu, W., Wu, G., Gao, G. F., Tan, W., & China Novel Coronavirus Investigating and Research Team (2020). A Novel Coronavirus from Patients with Pneumonia in China, 2019. The New England journal of medicine, 382(8), 727–733. https://doi.org/10.1056/NEJMoa2001017

2. Cucinotta, D., & Vanelli, M. (2020). WHO Declares COVID-19 a Pandemic. Acta Bio-MedicalJ: Atenei Parmensis, 91(1), 157–160. https://doi.org/10.23750/abm.v91i1.9397

3. WHO Coronavirus (COVID-19) Dashboard. (n.d.). With Vaccination Data. Retrieved January 9, 2023, from https://covid19.who.int/

4. Naqvi, A. A. T., Fatima, K., Mohammad, T., Fatima, U., Singh, I. K., Singh, A., Atif, S. M., Hariprasad, G., Hasan, G. M., & Hassan, M. I. (2020). Insights into SARS-CoV-2 genome, structure, evolution, pathogenesis and therapies: Structural genomics approach. Biochimica Et Biophysica Acta (BBA) -Molecular Basis of Disease, 1866(10), 165878.

5. Harvey, W. T., Carabelli, A. M., Jackson, B., Gupta, R. K., Thomson, E. C., Harrison, E. M., Ludden, C., Reeve, R., Rambaut, A., Peacock, S. J., & Robertson, D. L. (2021). SARS-CoV-2 variants, spike mutations, and immune escape. Nature Reviews Microbiology, 19(7), 409–424.

6. Tracking SARS-CoV-2 variants. (2022, October 27). https://www.who.int/activities/tracking-SARS-CoV-2-variants

7. G. La Rosa, M. Iaconelli, P. Mancini, G. Bonanno Ferraro, C. Veneri, L. Bonadonna, L. Lucentini, E. Suffredini First detection of SARS-CoV-2 in untreated wastewater in Italy. Sci. Total Environ., 736 (2020), p. 139652, https://doi.org/10.1016/j.scitotenv.2020.139652

8. A. Hata, H. Hara-Yamamura, Y. Meuchi, S. Imai, R. Honda. Detection of SARS-CoV-2 in wastewater in Japan during a COVID-19 outbreak. Sci. Total Environ. (2020), p. 143578, https://doi.org/10.1016/j.scitotenv.2020.143578

9. A. Nemudryi, A. Nemudraia, T. Wiegand, K. Surya, M. Buyukyoruk, C. Cicha, K.K. Vanderwood, R. Wilkinson, B. Wiedenheft Temporal detection and phylogenetic assessment of SARS-CoV-2 in municipal wastewater Cell Rep. Med., 1 (2020), p. 100098, https://doi.org/10.1016/j.xcrm.2020.100098

10. Yifan Zhu, Wakana Oishi, Chikako Maruo, Mayuko Saito, Rong Chen, Masaaki Kitajima, Daisuke Sano, Early warning of COVID-19 via wastewater-based epidemiology: potential and bottlenecks, Science of The Total Environment, Volume 767,2021,145124, ISSN 0048-9697, https://doi.org/10.1016/j.scitotenv.2021.145124

11. Oleksandr Panasiuk, Annelie Hedström, Jiri Marsalek, Richard M. Ashley, Maria Viklander, Contamination of stormwater by wastewater: A review of detection methods, Journal of Environmental Management, Volume 152, 2015, Pages 241–250, ISSN 0301-4797, https://doi.org/10.1016/j.jenvman.2015.01.050.

12. Wang, H., Liu, Q., Hu, J., Zhou, M., Yu, M. Q., Li, K. Y., Xu, D., Xiao, Y., Yang, J. Y., Lu, Y. J., Wang, F., Yin, P., & Xu, S. Y. (2020). Nasopharyngeal Swabs Are More Sensitive Than Oropharyngeal Swabs for COVID-19 Diagnosis and Monitoring the SARS-CoV-2 Load. Frontiers in medicine, 7, 334. https://doi.org/10.3389/fmed.2020.00334

13. Thomas S. Higgins, Arthur W. Wu, Jonathan Y. Ting, SARS-CoV-2 Nasopharyngeal Swab Testing False-Negative Results From a Pervasive Anatomical Misconception, JAMA Otolaryngology-Head & Neck Surgery November 2020 Volume 146; 993-994

14. Ministry of Science & Technology. PressReleasePage @ pib.gov.in. 2021. https://pib.gov.in/PressReleasePage.aspx?PRID=1722373 (accessed Oct 19, 2022)

15. Neeri’s gargling test kit gets DCGI’s nod. (2021, July 4). Times of India. Retrieved February 14, 2023, from https://www.google.com/amp/s/m.timesofindia.com/city/nagpur/neeris-gargling-test-kit-gets-dcgi-nod/amp_articleshow/84103217.cms

16. IHIP. Integrated Health Information Platform Integrated Disease Surveillance Programme Ministry of Health and Family Welfare, Government of India @ ihip.nhp.gov.in. https://ihip.nhp.gov.in/idsp/#!/.

17. Provisional Population Totals, Census of India 2011 Cities Having Population 1 Lakh and above Provisional Population Totals, Census of India 2011 Cities Having Population 1 Lakh and Above.; 2011.

18. WHO. Global Epidemiological Surveillance Standards for Influenza.; 2013. https://www.who.int/publications/i/item/9789241506601 Accessed on February 7, 2023

19. CSIR-NEERI_SARS-CoV-2 RT-PCR testing and WGS Protocol and facilities available for SARS-CoV-2.pdf. https://drive.google.com/file/d/1pMS_XBUwQ7QjCjwYscw6e0Ctw3Jjvsr0/view

20. Director CSIR-NEERI. (2021b, August 9). Saline Gargle RT PCR Innovation by CSIR NEERI [Video]. YouTube. https://www.youtube.com/watch?v=1OPZFlF4GpQ

21. Takara bio inc. RR037A For Research Use PrimeScript TM RT Reagent Kit (Perfect Real Time) Product Manual. https://www.takarabio.com/documents/UserManual/RR037A_e.v2008Da.pdf.

22. Oxford Nanopore Technologies. PCR Tiling of SARS-CoV-2 Virus with Rapid Barcoding and Midnight RT PCR Expansion (SQK-RBK110.96 and EXPMRT001). Vol 3.; 2021. https://nanoporetech.com/resource-centre/knowledge Accessed on February 7, 2023,

23. Wick RR, Judd LM, Holt KE. Performance of neural network base-calling tools for Oxford Nanopore sequencing. Genome Biol. 2019:1–10. https://doi.org/10.1186/s13059-019-1727-y

24. Commander, a command line free GUI-based sequencing data analysis tool developed by Genotypic Technology, Bangalore India https://www.genotypic.co.in/commander/. Accessed on February 7, 2023,

25. Toole ÁO, Scher E, Underwood A, et al. Assignment of epidemiological lineages in an emerging pandemic using the pangolin tool. 2021;7(2):1–9. https://doi.org/10.1093/ve/veab064

26. Outbreak.info SARS-COV-2 data explorer. outbreak.info. Retrieved March 22, 2023, from https://outbreak.info/

27. Wilhelm, A., Toptan, T., Pallas, C., Wolf, T., Goetsch, U., Gottschalk, R., Vehreschild, M. J. G. T., Ciesek, S., & Widera, M. (2021). Antibody-Mediated Neutralization of Authentic SARS-CoV-2 B.1.617 Variants Harboring L452R and T478K/E484Q. Viruses, 13(9), 1693. https://doi.org/10.3390/v13091693

28. Liu, Y., Liu, J., Johnson, B. A., Xia, H., Ku, Z., Schindewolf, C., Widen, S. G., An, Z., Weaver, S. C., Menachery, V. D., Xie, X., & Shi, P. Y. (2022). Delta spike P681R mutation enhances SARS-CoV-2 fitness over Alpha variant. Cell reports, 39(7), 110829. https://doi.org/10.1016/j.celrep.2022.110829

29. Kuzmina, A., Atari, N., Ottolenghi, A., Korovin, D., lass, I. C., Rosental, B., Rosenberg, E., Mandelboim, M., & Taube, R. (2022). P681 mutations within the polybasic motif of spike dictate fusogenicity and syncytia formation of SARS CoV-2 variants. BioRxiv, 2022.04.26.489630. https://doi.org/10.1101/2022.04.26.489630

30. Brito, A.F., Semenova, E., Dudas, G., et al. Global disparities in SARS-CoV-2 genomic surveillance. Nat Commun 13, 7003 (2022). https://doi.org/10.1038/s41467-022-33713-y

31. Chen, Z., Azman, A.S., Chen, X. et al. Global landscape of SARS-CoV-2 genomic surveillance and data sharing. Nat Genet 54, 499–507 (2022). https://doi.org/10.1038/s41588-022-01033-y

32. Genomic sequencing of SARS-CoV-2: a guide to implementation for maximum impact on public health. Geneva: World Health Organization; 2021. License: CC BY-NC-SA 3.0 IGO.

33. Schrader, C., Schielke, A., Ellerbroek, L., & Johne, R. (2012). PCR inhibitors - occurrence, properties, and removal. Journal of applied microbiology, 113(5), 1014–1026. https://doi.org/10.1111/j.1365-2672.2012.05384.x

34. Bickley, J., Short, J. K., McDowell, D. G., & Parkes, H. C. (1996). Polymerase chain reaction (PCR) detection of Listeria monocytogenes in diluted milk and reversal of PCR inhibition caused by calcium ions. Letters in applied microbiology, 22(2), 153–158. https://doi.org/10.1111/j.1472-765x.1996.tb01131.x

35. Kirkland, P. D., & Frost, M. J. (2020). The impact of viral transport media on PCR assay results for the detection of nucleic acid from SARS-CoV-2. Pathology, 52(7), 811–814. https://doi.org/10.1016/j.pathol.2020.09.013

36. Rural population (% of total population) - India. data.worldbank.org. Retrieved February28,2023,from https://data.worldbank.org/indicator/SP.RUR.TOTL.ZS?locations=IN

37. Priyadarshini, S. (2021, May 21). India’s sewage surveillance for SARS-CoV-2 going down the drain. Nature India; Nature Portfolio. https://doi.org/10.1038/nindia.2021.75

38. Kandel, C. E., Young, M., Serbanescu, M. A., Powis, J. E., Bulir, D., Callahan, J., Katz, K., McCready, J., Racher, H., Sheldrake, E., Quon, D., Vojdani, O. K., McGeer, A., Goneau, L. W., & Vermeiren, C. (2021). Detection of severe acute respiratory coronavirus virus 2 (SARS-CoV-2) in outpatients: A multicenter comparison of self-collected saline gargle, oral swab, and combined oral-anterior nasal swab to a provider collected nasopharyngeal swab. Infection control and hospital epidemiology, 42(11), 1340–1344. https://doi.org/10.1017/ice.2021.2

39. Goldfarb, D. M., Tilley, P., Al-Rawahi, G. N., Srigley, J. A., Ford, G., Pedersen, H., Pabbi, A., Hannam-Clark, S., Charles, M., Dittrick, M., Gadkar, V. J., Pernica, J. M., & Hoang, L. M. N. (2021). Self-Collected Saline Gargle Samples as an Alternative to Health Care Worker-Collected Nasopharyngeal Swabs for COVID-19 Diagnosis in Outpatients. Journal of clinical microbiology, 59(4), e02427–20. https://doi.org/10.1128/JCM.02427-20

40. Saito, M., Adachi, E., Yamayoshi, S., Koga, M., Iwatsuki-Horimoto, K., Kawaoka, Y., & Yotsuyanagi, H. (2020). Gargle Lavage as a Safe and Sensitive Alternative to Swab Samples to Diagnose COVID-19: A Case Report in Japan. Clinical infectious diseases: an official publication of the Infectious Diseases Society of America, 71(15), 893–894. https://doi.org/10.1093/cid/ciaa377

41. Bennett, S., Davidson, R. S., & Gunson, R. N. (2017). Comparison of gargle samples and throat swab samples for the detection of respiratory pathogens. Journal of virological methods, 248, 83–86. https://doi.org/10.1016/j.jviromet.2017.06.010

