## Supplementary Figure for "Non-invasive SARS-CoV-2 genome surveillance and its utility in resource-poor settings during the Delta wave of the COVID-19 pandemic"

### SARS-CoV-2 major variant pattern

DISTRIBUTION OF SARS-CoV-2 OTHER VARIANTS IN GARGLE BASED GENOMIC SURVEILLANCE FROM MARCH - DECEMBER 2021

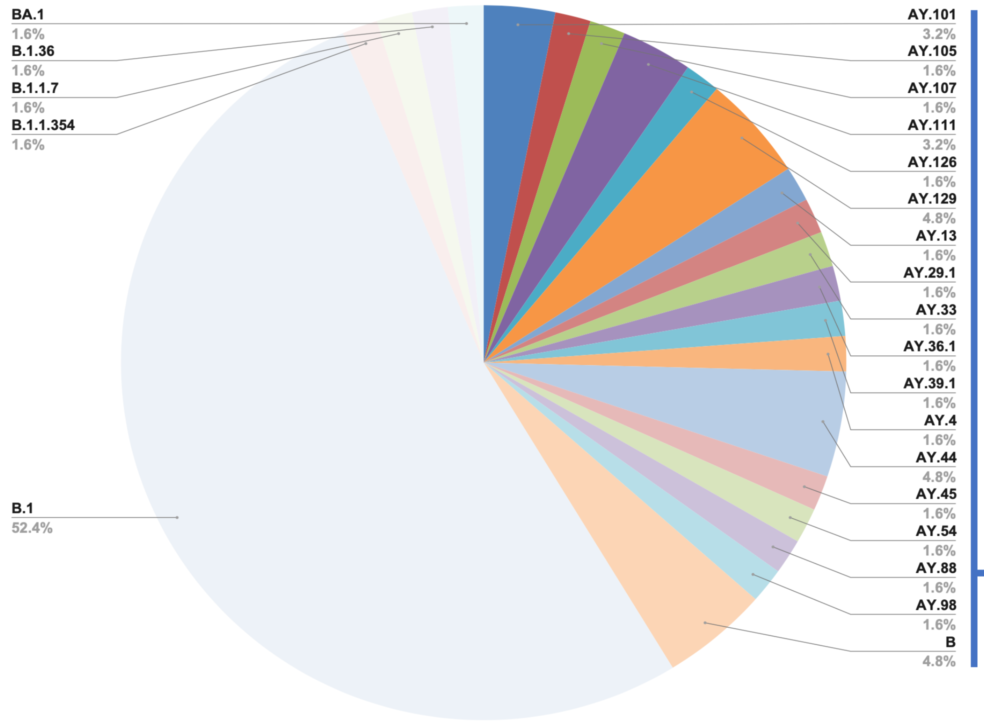


DISTRIBUTION OF SARS-CoV-2 MAJOR VARIANTS IN GARGLE BASED GENOMIC SURVEILLANCE FROM MARCH - DECEMBER 2021

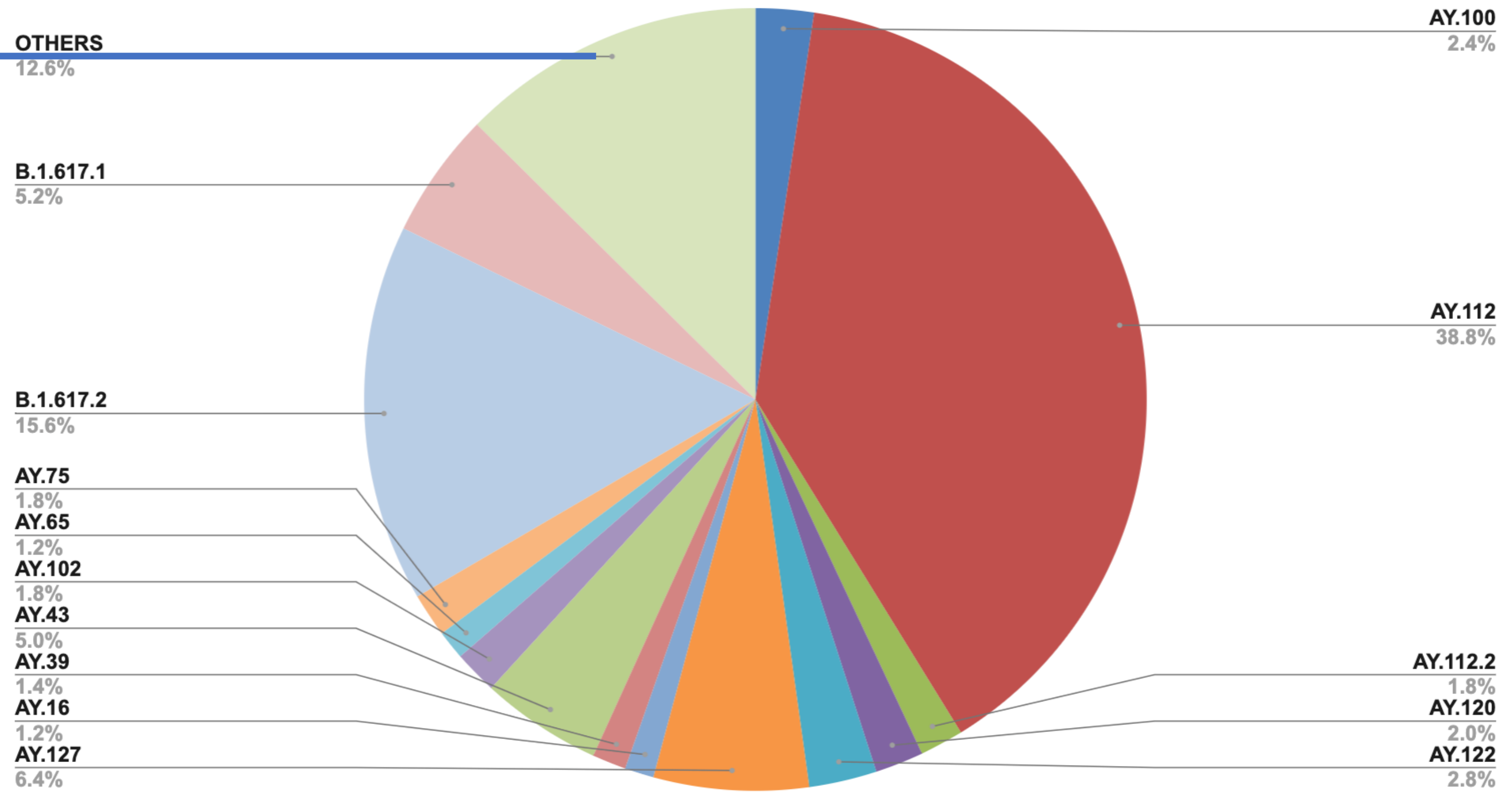
